# Impact of bariatric surgery on multiple health outcomes: a protocol for umbrella review of systematic review and meta-analysis

**DOI:** 10.1101/2020.08.22.20180018

**Authors:** Min Seo Kim, Jong Yeob Kim, Sungsoo Park

**Affiliations:** Korea University College of Medicine, Seoul, Republic of Korea.; Cheongsan Public Health Center, Wando, Ministry of Health and Welfare, Republic of Korea.; Yonsei University College of Medicine, Seoul, Republic of Korea.; Division of Upper Gastrointestinal Surgery, Department of Surgery, Korea University College of Medicine, Seoul, Republic of Korea

## Abstract

The number of bariatric surgery rises as the prevalence of obesity and metabolic comorbidities consistently increases[1]. Although bariatric surgery was originally developed for glycemic control and weight reduction, increasing evidence suggested extra-metabolic health outcomes are followed by bariatric surgery; incidences on diverse types of cancer[2], perinatal outcomes[3], sexual function[4], and even degree of physical activity[5] are known to be altered after bariatric surgery. We aim to conduct umbrella review for metabolic and other multiple health outcomes following bariatric surgery, and systematically appraise the context and quality of the relevant evidence.

## METHODS

This investigation is currently under review for registration on the International Prospective Register of Systematic Reviews (PROSPERO). If deviations are made to the prospectively registered protocol, they will be reported in the supplementary material of final publication.

### Eligibility Criteria

#### Condition or domain being studied

Health outcomes of interest include metabolic outcomes (weight/glycemic control), other outcomes (e.g., incidences and courses of multiple cancers, fracture, mental health, psychiatric/cognitive disorders, cardiovascular diseases, gastroenterological diseases, rheumatoid diseases, respiratory diseases, renal functions, urologic disorders, etc.), and biomarkers (e.g., ghrelin, uric acid, GLP-1, PYY, GIP, Gut hormones, serum inflammatory markers, adiponectin, chemerin, plasminogen activator inhibitor-1, leptin, resistin, visfatin, etc.)

#### Participants/population

For the analysis of the incidence of multiple health outcomes, patients who have not been diagnosed with such health consequences prior to initiation of the study or have recorded disease-free status will be included. For the analysis of the disease course/progression/functional change of multiple health outcomes, patients diagnosed with predefined health outcomes will be included in analysis. Incidence and clinical course of multiple health outcomes will be analyzed separately as they involve different inclusion criteria and thereby have discrete casemixes.

#### Interventions/exposures

The intervention of interest is a BS which includes Roux-en Y gastric bypass (RYGB), sleeve gastrectomy (SG), biliopancreatic diversion (BPD), biliopancreatic diversion with or without duodenal switch (BPD/DS), and gastric banding. Biliopancreatic diversion (BPD) and biliopancreatic diversion with duodenal switch (BPD-DS) will be assumed as a single surgical technique as described in previous studies[6, 7].

#### Comparator/control

Comparators will be either pre-surgical status of surgical patients or control group comprised of patients who have not undergone BS (healthy patients or patients subject to lifestyle modification or medical treatment).

#### Types of Studies (inclusion and exclusion criteria)

Studies of the following forms will be included:

- meta-analyses of randomized controlled trials (RCT) or observational studies.
- systematic reviews with quantitative synthesis.
- meta-analyses presenting results of pre-versus post-BS or surgical patients versus non-surgical controls.
- studies investigated on the general population.
- studies presenting clinical outcomes from equal to or longer than mean of 12 months follow-up duration (The restriction on follow up period will be exclusively applied for studies investigating metabolic outcomes of BS, and not for other outcomes.)

Studies of the following forms will be excluded:

- literature review with only qualitative synthesis and/or theoretical explanation.
- opinion pieces or comments.
- meta-analyses with insufficient data for analyses.
- systematic reviews without meta-analyses.
- studies investigating revisional bariatric surgery, reoperation, or surgical failure.
- studies exploring endoscopic techniques such as intragastric balloon placement and endoscopic sleeve gastroplasty for intentional weight loss.
- Studies focusing on efficiency of modified technique of conventional bariatric surgery (e.g., single incision, one-anastomosis, vertical sleeve, hand-sewn versus mechanical gastrojejunal anastomosis, staple technique, etc.)
- Studies focusing on efficiency of endoscopic techniques intended to reduce body weight such as intragastric balloon placement and endoscopic sleeve gastroplasty were excluded as they are perceived as procedures rather than surgery.
- studies focusing on perioperative outcomes, postoperative complications, or acute consequences such as a change in biomarker within postop 7days after bariatric surgery.
- studies comparing different BS techniques (e.g., RYGB versus sleeve gastrectomy, etc.)
- studies investigating the predictive/prognostic value of bariatric surgery.
- studies comparing different surgical modalities of BS (e.g., open versus laparoscopic versus robotic BS).
- studies exclusively investigating specific populations and thus unable to extrapolate the results to general populations (e.g., child/adolescent, elderly, black, Asian, etc.)
- studies involving animal experiments or in vitro results.

### Primary Outcomes

The main outcomes will be the incidences of multiple health outcomes after BS. Measures of the effect of BS on health outcomes can be presented as metrices such as odds ratio (OR), relative risk (RR), and hazard ratio (HR). We will use outcome metrices identical to those reported in the original meta-analyses.

### Secondary Outcomes

The secondary outcomes will be clinical courses/outcomes, improvement/aggravation, and functional change of multiple health outcomes after BS. Measures of the effect of BS on health outcomes can be presented as metrices such as odds ratio (OR), relative risk (RR), hazard ratio (HR), mean difference (MD), standardized mean difference (SMD), and weighted mean difference (WMD). We will use outcome metrices identical to those reported in the original meta-analyses.

### Search Methods for Identification of Studies

#### Electronic Database Search

We will search on PubMed, Embase, and The Cochrane Library (Cochrane database of systematic reviews) from inception to July 2020. Two researchers (MS Kim and JY Kim) will independently search for systematic reviews and meta-analyses investigating the effect of bariatric surgery (BS) on multiple health outcomes.

##### PUBMED

(“bariatric*”[tiab] OR “sleeve gastrectom*”[tiab] OR “gastric sleeve*”[tiab] OR “roux-en-Y”[tiab]

OR “gastric bypass*”[tiab] OR “RYGB”[tiab] OR “gastric band*”[tiab] OR “diversion*”[tiab]

OR “Bariatric Surgery”[Mesh] OR “Anastomosis, Roux-en-Y”[Mesh] OR “Gastric Bypass” [Mesh])

AND (meta-analy*[tiab] OR “meta-analysis”[pt])

NOT protocol[ti] NOT comment*[ti]

##### EMBASE

(bariatric*:ab,ti OR ‘sleeve gastrectom*’:ab,ti OR ‘gastric sleeve*’:ab,ti OR ‘roux en y’:ab,ti OR ‘RYGB’:ab,ti OR ‘gastric band*’:ab,ti OR ‘diversion*’:ab,ti OR

‘bariatric surgery’/exp OR ‘sleeve gastrectomy’/exp OR ‘gastric sleeve’/exp OR ‘roux-en-y gastric bypass’/exp OR ‘gastric banding’/exp)

AND (‘meta analy*’:ti)

NOT (‘conference abstract’:it OR ‘conference paper’:it OR ‘conference review’:it OR editorial:it OR note:it OR letter:it OR ‘short survey’:it)

##### CDSR

“Title Abstract Keyword”

(“bariatric*” OR “sleeve gastrectom*” OR “gastric sleeve*” OR “roux-en-Y” OR “gastric bypass*” OR “RYGB” OR “gastric band*” OR “diversion*”)

### Data Collection and Analysis

#### Data extraction (selection and coding)

Two researchers (JY Kim and MS Kim) will independently search the existing literature using a predetermined search strategy and also manually search through reference lists of the articles found through the initial search. Studies that meet inclusion criteria will be gathered, and duplicates will be removed. Titles, abstracts, and full text of each study will be reviewed for inclusion. Ambiguous studies will be managed through discussion and agreement between two authors. If the discrepancy between the two authors persists, a third party (S Park, corresponding author) will make the decision. The study selection process will be recorded using PRISMA flowchart (David et al, 2009).

A predefined data extraction table will be used to collect data and summarize each study. Following details will be obtained: publication year, number of included studies in a meta-analysis, number of events and patients, the model of effect estimation (random effects or fixed effects), heterogeneity, outcome of interest (including metabolic and secondary outcomes), and evidence level (GRADE, etc.).

When extracting pooled effect sizes from included studies, we prefer to use pooled effect sizes from studies that present details of all individual studies (e.g., presenting forest plot) over studies that only demonstrate summary pooled effect sizes.

#### Risk of Bias (quality) assessment

The methodological quality of included meta-analyses will be assessed using the validated AMSTAR 2 (A Measurement Tool to Assess Systematic Reviews 2) instrument. We will not examine the quality of the individual cohort studies included in the meta-analyses as this work is conducted by the authors. The certainty of evidence for each main/primary outcome will be evaluated with GRADE (Grading of Recommendations Assessment, Development and Evaluation) approach, as has been done in numerous previous umbrella reviews[8–12]. The certainty of evidence will be classified as high, moderate, low or very low. Small study effect will be assessed with Egger’s test of funnel plot asymmetry.

#### Strategy for data synthesis

We will re-analyze each extracted meta-analysis under both fixed and random effects model, adhere to the workflow for pairwise meta-analyses described elsewhere[13, 14]. Pooled effect sizes and 95% confidential intervals (CI) will be used for the re-analysis. We will use the same metrices as used in the original meta-analyses (RR, OR, HR, MD, SMD, WMD). If included studies (meta-analyses) provided confounder-adjusted effect sizes, we will preferentially use the adjusted effect sizes. Heterogeneity between studies will be calculated as I^2^. Egger’s p value and prediction interval will also be calculated. Further details of the methodology for the bias analyses are demonstrated in previous works[15, 16]. The results of the primary and significant outcomes (effect size and evidence level (GRADE)) will be used to construct an evidence map. Software R and its packages will be used for the analysis.

#### Subgroup and Sensitivity Analyses

Subgroup/subset analyses will be performed when available, by these factors:

- different surgical techniques (RYGB, SG, BPD, BPD-BS, etc) for metabolic outcomes
- study design (randomized controlled trials and observational studies)
- Subset analysis of studies which properly controlled for baseline BMI of experimental and control groups, such as RCTs, pre-versus post-BS studies, and observational studies using BMI-matched control group

Outcomes from subgroup/subset analyses are not subject to bias analyses (e.g., Egger’s test, heterogeneity test, 95% prediction interval, etc) and therefore will not be included in the evidence map/evidence level stratification.

## Data Availability

Contact to

## ACKNOWLEDGEMENTS

None.

## AUTHOR STATEMENT

None

## CONFLICTS OF INTEREST

The authors declare no potential conflicts of interest.

## AUTHOR APPROVAL

All authors have been seen and approved the protocol

## Notes

### Competing Interest Statement

The authors have declared no competing interest.

